# Diagnostic concordance between physiotherapist and emergency physicians for patients with a musculoskeletal disorder in the emergency department

**DOI:** 10.1101/2020.10.28.20221762

**Authors:** Rose Gagnon, Kadija Perreault, Gabrielle Brun, Eveline Matifat, François Desmeules, Simon Berthelot, Luc J. Hébert

## Abstract

**Question:** What is the level of diagnostic concordance between physicians and physiotherapists for patients consulting in an emergency department (ED) with musculoskeletal disorders (MSKDs)?

**Design:** Secondary analysis of data obtained through a pragmatic randomized controlled trial, physiotherapist (PT) and emergency physicians (EPs) unblinded, per-protocol analysis.

**Participants:** 78 participants aged 18 to 80 years presenting with a minor MSKD were recruited and data from the 40 participants randomized in the intervention group were used (mean age 36.6 yrs (SD: 17.3, IQR: 22.0; 46.8); women: 55%).

**Outcome measures:** Diagnostic concordance was established using the International Classification of Diseases 11 (ICD-11) and examined between EP and PT using raw agreement and Gwet’s first-order agreement coefficient (AC1).

**Results:** Forty participants were recruited and 36 were assessed by the PT and EP (36.8 ± 18.2 years old; 55.6% women). Overall raw agreement was 86.1% and an almost perfect diagnostic concordance was observed between PT and EP (Gwet’s AC1: 0.84, 95% CI: 0.69 – 0.98). The most common reason for disagreement was suspected bone fracture or contusion in comparison with ligament or meniscus disorder.

**Conclusion:** Results from this study indicate that overall diagnostic concordance between PT and EP in the ED for patients consulting with MSKDs is almost perfect. These results tend to support the safety of such models of care. However, more studies are needed to confirm these results as the EP wasn’t always independent and the classification used was imprecise.

**Registration:** #NCT04009369

## INTRODUCTION

Overcrowding and long waits in the emergency department (ED) have been worrisome problems for several years in many hospitals around the world. Overcrowding in the ED is thought to lead to several adverse effects such as increased time to care, increased length of stay in the ED, and increased medical errors. ^1–5^ One of the main causes of the increase in the number of ED visits is the unavailability of primary care services. ^6,7^ As a response, many patients turn to the ED for the management of their condition in such a context, ^8^ especially when suffering from musculoskeletal disorders (MSKDs) which are among the most frequent presenting complaints, representing between 18% and 25% of all visits in some EDs. ^9–11^ Patients who consult for MSKDs are often assigned a low priority at ED triage due to the non-urgent nature of their condition. ^12^ As a consequence of ED overcrowding, hospitals in several countries have integrated, after initial nurse triage, direct access and autonomous management by physiotherapists (PT) of MSKDS, often labelled as advanced practice PTs (APPs). Studies conducted in the United Kingdom and in Australia indicate that initial assessment by a PT in the ED rather than by an emergency physician (EP) for MSKDs could reduce pain and disability, the average time to care, the average length of stay in the ED, the number of redundant medical consultations, the number of unnecessary drug prescriptions and imaging tests, an thus optimize overall management. ^9,13–15^

Although the current literature on the integration of a PT in different clinical contexts such as EDs or orthopaedic clinics demonstrates potential benefits, few studies have examined the diagnostic concordance between PTs and physicians. ^16^ While ways of reporting diagnostic concordance vary between studies, some studies carried out in orthopaedic clinics have reported proportions of diagnostic agreement between PTs and orthopaedic surgeons ranging from 74.5% to 90% and Kappa coefficients ranging from 0.69 to 0.70. ^16^ No significant differences were observed in terms of diagnostic accuracy between the two professionals in regards to arthroscopy or MRI. ^16^ Regarding the ED, to our knowledge, only one previous study looked specifically at the diagnostic concordance between a PT and an EP in the ED. In a study conducted in 2019 (n=113), Matifat et al. reported a raw agreement between the two professionals of 87% and a Kappa coefficient of 0.81 (95% CI: 0.72-0.90). ^17^ However, the agreement was lower in regards to treatment modalities to be used and recommended discharge plan. Moreover, in a study by Sutton et al., a PT in the ED managed more than 1300 patients without any misdiagnoses or adverse events (95% CI: 0.0-0.3%). ^18^ A better understanding of diagnostic concordance between the PT and the EP in the ED could supportdecisions regarding the implementation of such models of care.

Therefore, to pursue in this direction and to contribute to the adding of knowledge on diagnostic concordance, the research question for our study was, in patients presenting to the ED with a MSKD,

1. What is the level of diagnostic concordance between EP and PT?

## METHOD

### Design

This study was a secondary analysis of data obtained through a pragmatic randomized clinical trial that was conducted in a major academic ED. [REF] Two groups were recruited over a 24-week period from September 2018 to March 2019. The intervention group had direct access to a PT in the ED and the control group was managed in the usual way by an EP. Participants were randomized using block randomization with stratification by affected body region. This study was approved by the Research Ethics Committee of the CHU de Québec – Université Laval and registered at the US National Institutes of Health #NCT04009369.

### Participants, therapists, centres

To be considered eligible, patients presenting to the ED had to meet the following criteria: 1) be aged between 18 and 80 years, 2) have a minor peripheral or vertebral MSKD with or without trauma, 2) have a triage score of 3 (urgent), 4 (less urgent), or 5 (non-urgent) according to the Canadian Emergency Department Triage and Acuity Scale classification ^19^, 3) have the ability to legally consent to participate, 4) understand French in order to complete the study questionnaires orally or in writing, and 5) be a beneficiary of the provincial health insurance plan (*Régie de l’assurance maladie du Québec*). Patients were not considered eligible if they had one of the following criteria: 1) a MSKD requiring emergent care (e.g. open fracture, dislocation, open wound), 2) a red flag (e.g. progressive neurological disorder, infectious symptoms), 3) a concomitant unstable clinical condition (e.g. pulmonary, cardiac, digestive and/or psychiatric), or 4) if they were already hospitalized or living in a long-term care facility.

### Intervention

Once the eligible patients had been triaged and their information registered into the CHUL’s electronic information system, the research coordinator (RG) met with them to explain the project, answer their questions, and obtain their informed consent. Each participant was then instructed to complete a questionnaire in which they were asked to provide socio-demographic information such as age, gender, presence of self-reported other health conditions and income level. Thereafter, they were assigned to one of the two groups according to the randomization sequence previously stated. Each participant assigned to the intervention group was managed by a PT. Following his interview, the PT conducted a brief physical examination of each participant. The PT was then asked to complete a standardized form detailing diagnosis and recommended interventions. He also completed his usual clinical note in the patient’s medical record. The standardized form and a copy of his clinical note were then included in the consultation request submitted to the EP. The ongoing internal policy at the CHU de Québec – Université Laval during our study was to have all participants in the intervention group seen by the EP following the PT’s assessment, thereby preventing both assessors from being independent. Given the pragmatic nature of this study and the current legislative context, the EP had access to the assessment and diagnosis of the PT, but it is impossible to know whether the EP consulted them in a systematic manner or not. In addition, the professionals were free to communicate with each other regarding the management of individual patients. The diagnosis made by the EP were extracted from each participant’s electronic records after their ED stay.

### Outcome measures

#### Primary outcome

Our primary outcome was the diagnosis made by the PT and the EP for all participants in the intervention group. Since the vocabulary used by each professional for the same condition can sometimes differ, we first used the International Classification of Diseases 11 (ICD-11) as the basis for our analyses. ^20^ Hence, the diagnoses made by each professional were associated with the corresponding ICD-11 diagnostic code. We then identified the higher order diagnostic categories associated with the initial diagnostic codes, again based on the ICD-11. The diagnostic categories were as follows: disorder of ligament or meniscus (e.g. sprain), soft tissue disorder (muscle, fascia, tendon, cartilage), fracture or contusion, dizziness and giddiness, conditions associated with the spine, and overuse or chronic pain (e.g. osteoarthritis, recurrent instability). The assignment of ICD-11 diagnostic codes, grouping into higher order diagnostic categories, and comparison of diagnoses was performed by two independent assessors (RG and GB). A third evaluator was asked to settle on the concordance in case of any disagreement (LJH, this step was used for 3 cases).

### Data analysis

Participants characteristics were calculated using descriptive statistics. For the concordance analyses, we considered the primary diagnosis made by both providers. We examined the overall diagnostic concordance using the raw agreement and Gwet’s first-order agreement coefficient (AC1) (R Software, 4.0.2; package irrCAC, 1.0, proc gwet.ac1.raw) and reported a corresponding 95% confidence interval (CI). In addition to being less affected by prevalence and marginal probability than Cohen’s Kappa, Gwet’s AC1 has been shown to have a more stable inter-rater reliability coefficient than Cohen’s Kappa. ^21^ We interpreted the strength of agreement between the two professionals as proposed by Landis and Koch and reported by Sim and Wright. according to the following scale: ≤ 0 = poor, .01-.20 = slight, .21-0.40 = fair, .41-.60 = moderate, .61-0.80 = substantial et .81-1 = almost perfect. ^22^

## RESULTS

### Flow of participants, therapists, centres through the study

589 participants were assessed for eligibility between September 2018 and March 2019 and 78 of them were recruited. The reasons for exclusion and the demographic characteristics of all participants are presented in the randomized controlled trial article. [REF] The demographic characteristics of the participants managed by the PT are presented in Table 1. Participants’ mean age was 36.8 years (SD: 18.2), and 55.6% were female. The mean pain intensity was 6.9 on a 0-10 scale (SD: 2.2) and the mean pain interference on function was 4.2 out of 10 (SD: 2.2). Moreover, participants had a mean pain catastrophizing score of 18.5 (/100) (SD: 13.3).

**Table 1.** Demographic characteristics (n=36)

| Characteristics | PT (n=36) | IQR | 95% CI |
| --- | --- | --- | --- |
| Age (yrs), mean (SD) | 36.8 (18.2) | 22.0 - 48.5 |  |
| Gender, n males (%) | 16 (44.4) |  | 28.3 – 61.7 |
| Triage category in ED, n (%) |  |  |  |
| Urgent (P3) | 14 (38.9) |  | 23.6 – 56.5 |
| Semi-urgent (P4) | 22 (61.1) |  | 43.5 – 76.4 |
| Non-urgent (P5) | 0 (0) |  |  |
| Living arrangements, n (%) |  |  |  |
| Alone | 10 (27.8) |  | 14.8 – 45.4 |
| With family/Spouse | 26 (72.2) |  | 54.6 – 85.2 |
| Occupation, n (%) |  |  |  |
| Medical leave due to present complaint | 3 (8.3) |  | 2.2 – 23.6 |
| Medical leave for other reason | 0 (0) |  |  |
| Working Full time | 15 (41.7) |  | 26.0 – 59.1 |
| Working Part time | 1 (2.8) |  | 0.1 – 16.2 |
| Student | 10 (27.8) |  | 14.8 – 45.4 |
| Other | 7 (19.4) |  | 8.8 – 36.6 |
| Loss of wages since onset, yes n (%) | 3 (8.3) |  | 2.2 – 23.6 |
| Personal income, n (%) |  |  |  |
| 1\$ - 49 999\$ | 23 (71.9) | | 53.0 – 85.6 |
| 50 000\$ - 99 999\$ | 9 (28.1) | | 14.4 – 47.0 |
| ≥ 100 000\$ | 0 (0) | | |
| Highest level of education completed, n (%) |  |  |  |
| Primary or High School | 4 (11.4) |  | 3.7 – 27.7 |
| College or University | 31 (88.6) |  | 72.3 – 96.3 |
| Time since onset of symptoms, n weeks (%) |  |  |  |
| 0-2 | 27 (79.4) |  | 59.4 – 89.0 |
| 3-12 | 3 (8.8) |  | 2.2 – 24.2 |
| >12 | 4 (11.8) |  | 3.7 – 27.7 |
| Don't know | 1 (2.9) |  | 0.1 – 16.6 |
| Other health conditions, yes n (%) | 13 (36.1) |  | 21.3 – 53.8 |
| Pain level /10, mean (SD) | 6.9 (2.2) | 6.0 – 8.0 |  |
| Pain interference on function /10, mean (SD) | 4.2 (2.2) | 3.0 – 5.8 |  |
| Pain catastrophizing, mean (SD) | 18.5 (13.3) | 8.0 – 29.8 |  |

PT: intervention group, Yrs: years, ED: Emergency department

PT in the ED and related diagnoses were provided by a single senior PT with more than 10 years of experience with a variety of clienteles such as various MSKDs, geriatrics, and chronic pain.

Our study was conducted at the CHU de Québec – Université Laval, more precisely at the Centre hospitalier de l’Université Laval (CHUL), an academic ED located in Quebec City (Canada). More than 68 000 patients visited the CHUL ED in 2015, including more than 10 000 who did so for a MSKD.

### Compliance with trial method

Forty participants were assigned to the intervention group and 38 to the control group. Four participants were managed by the direct access PT, but left the ED before being managed by the EP. Therefore, only 36 participants were included in the concordance analyses.

### Diagnostic concordance between PT and EP

Overall, raw agreement between providers was 86.1% and the Gwet’s AC1 was 0.84 (95% CI: 0.69 – 0.98, p<.0001) (Table 2). The matrix of concordance between the PT and the EP is presented in Figure 2. The most common reason for disagreement was the PT suspecting a bone fracture or contusion while the EP’s diagnosis was a ligament or meniscus disorder. Two participants were diagnosed with a disorder of ligament or meniscus by the PT and a soft tissue disorder by the EP.

**Table 2.** Diagnostic concordance between emergency physicians and a direct access physiotherapist for patients presenting with a MSKD in the ED (n=36)

|  | Agreement by chance | Raw agreement | Gwet's AC1 | 95% CI | *p* |
| --- | --- | --- | --- | --- | --- |
| Diagnostic concordance | 15.8% | 86.1% | 0.84 | 0.69-0.98 | < .0001 |

Agreement by chance: Proportion of agreement due to raters agreeing on a rating, despite the fact that one or all of them may have given a random value <sup>21</sup>

No adverse event was reported during the study in either group.

## DISCUSSION

The aim of our study was to answer the following research question: In patients presenting to the ED for a MSKD, what is the level of diagnostic concordance between EP and PT? Overall, we observed an almost perfect diagnostic concordance between the PT and the EP.

While the level of diagnostic concordance obtained in our study is considered almost perfect, a few minor disagreements between the two professionals were observed. Indeed, on three occasions, the PT judged that the participant had a fracture or contusion, whereas the EP’s diagnosis was a ligament or meniscus disorder. These can be largely explained by the fact that, at the time of our study, the PT was not legally allowed to independently prescribe imaging tests. In some contexts, PTs using an advanced practice can autonomously prescribe and summarily interpret some imaging exams such as plain radiographs following intensive training. ^23^ In our study, the PT had to make a diagnosis based only on clinical signs and symptoms that could sometimes suggest a fracture and not just a sprain. The EP had access to any imaging tests he deemed necessary, allowing him to quickly confirm or rule out the presence of a fracture on a plain radiograph. Thus, although there was some level of discordance between the two professionals, it is possible that the level of diagnostic agreement could have been even higher if the PT would have been allowed to independently prescribe and interpret a plain radiograph. However, the PT was still able to identify all of the fractures in the intervention group without the use of imaging tests.

Our study is the second study addressing diagnostic concordance between a PT and EP in the ED. The study of diagnostic concordance is important to support the development of models of interdisciplinary collaboration in the ED. Indeed, some patients and specialists may not always feel comfortable with a collaborative care model because they feel that the care given by another professional might not always be of the same quality. ^24^ Near perfect diagnostic concordance is related to the safety of having a PT in the ED for patients with a variety of MSKDs. Moreover, these results highlight the potential role of various professionals such as the PT in developing new and potentially more effective trajectories of care in the ED for MSKD patients. ^25^

There is currently no consensus definition of diagnostic concordance in the literature. Studies that address this concept often take place in a variety of settings such as orthopaedics clinics or EDs and include various professionals with varying levels of expertise. It can therefore be arduous to compare results between them. ^17,25,26^ Moreover, diagnostic concordance analyses oftentimes do not allow to consider the level of diagnostic precision provided by each professional. Results from this study must therefore be interpreted with caution.

Although our sample size is small, the participants in the study presented a variety of MSKDs as previously reported. [REF] A major limitation of our study is that the EP had the possibility of knowing the PT’s diagnosis before making his own assessment and was therefore not blinded. However, our study used a pragmatic design and hence assessed real-life practice where the physicians have access to the PT’s files. Furthermore, the results presented in our study are very similar to those obtained by Matifat et al. in which study the PT and the EP were independent. It is thus possible that this limitation had a minimal impact on the diagnostic concordance level obtained. Finally, some diagnostic categories based on the ICD-11 such as “Conditions associated with the spine” were very imprecise, which may have had an upward impact on diagnostic concordance.

The results of this study suggest that the overall diagnostic concordance between PT and EP in the ED is almost perfect. Additional multicenter studies with larger sample sizes are needed to verify the external validity of these results. In addition, it would also be interesting to conduct further studies where the PT would be allowed to prescribe imaging tests such as plain radiograph to observe its impact on the level of agreement between physicians and PTs in the context of an ED.

**Figure 1.**
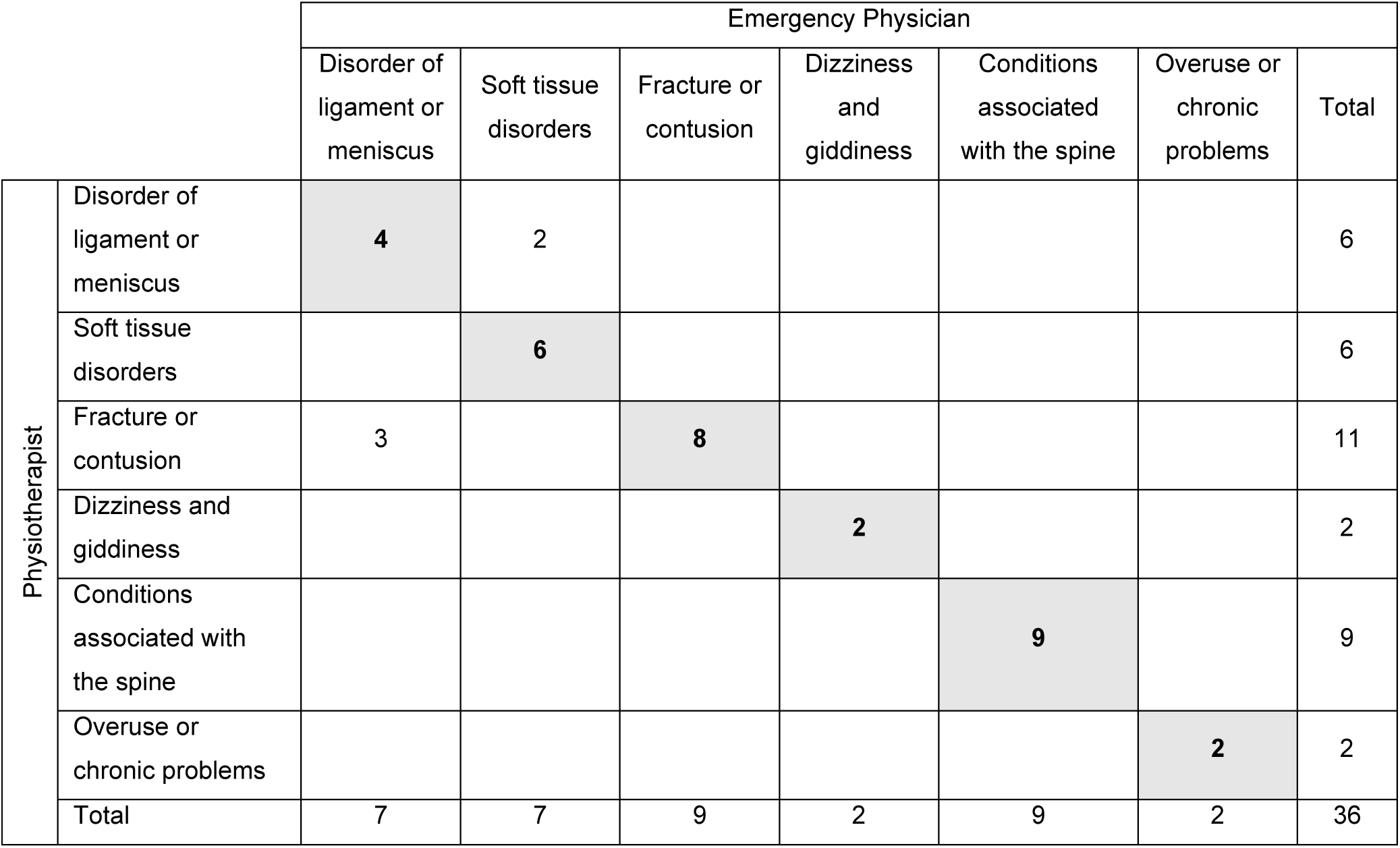
Matrix of diagnostic concordance between the physiotherapist and emergency physicians based on the ICD-11

## Data Availability

The data that support the findings of this study are available from the corresponding author, LJH, upon reasonable request.

## Acknowledgements

*We would like to thank the following persons for their contribution to project implementation* :

*All project participants, Antony Barabé PT, physiotherapist at the Centre Hospitalier de l’Université Laval (CHUL), the entire team of managers at the* Direction des services multidisciplinaires *of the CHU de Québec – Université Laval (Marie-Christine Laroche, Catherine van Neste, Marie-Claude Brodeur and Stéphane Tremblay) for their support throughout the implementation of the project and its realization, and Mathieu Blanchet, MD, FRCPC, head of the CHUL ED department during the duration of the study*.

## Notes

**Source(s) of support:** Support for this project was provided by the CHU de Québec – Université Laval, subsidies from LJH and KP and a research grant awarded by the multidisciplinary council of the CHU de Québec – Université Laval in association with the Fondation du CHU de Québec. RG received scholarships from the Canadian Institutes of Health Research, the CIRRIS, the Ordre professionnel de la physiothérapie du Québec and the Department of Rehabilitation funds of the Faculty of Medicine at Université Laval. KP is a Junior 1 Research Scholar from the Fonds de recherche du Québec-Santé (FRSQ).

### Competing Interest Statement

The authors have declared no competing interest.

### Clinical Trial

NCT04009369

### Funding Statement

Funding for this project was provided by the CHU de Québec-Université Laval, subsidies from LJH and KP and a research grant awarded by the multidisciplinary council of the CHU de Québec-Université Laval in association with the Fondation du CHU de Québec. RG received scholarships from the Canadian Institutes of Health Research, the CIRRIS, the Ordre professionnel de la physiothérapie du Québec and the Department of Rehabilitation funds of the Faculty of Medicine at Université Laval. KP is a Fonds de recherche du Québec-Santé (FRSQ) Junior 1 Research Scholar.

### Author Declarations

The CHU de Québec-Université Laval Ethics Committee approved this study. All participants gave written informed consent before data collection began.

